# Hippocampal Asymmetry Captures Non–Amyloid-Related Risk of Memory Decline and Clinical Progression

**DOI:** 10.64898/2026.01.06.26343553

**Authors:** Elham Ghanbarian, Babak Khorsand, Lukai Zheng, Davis C. Woodworth, Crystal M. Glover, Maria M. Corrada, Joshua D. Grill, S. Ahmad Sajjadi, the Alzheimer’s Disease Neuroimaging Initiative, Ali Ezzati

## Abstract

**Background:** Hippocampal atrophy is a key marker of Alzheimer’s disease (AD)-related neurodegeneration; however, hippocampal volume alone may not fully capture heterogeneity in cognitive decline. Hemispheric hippocampal asymmetry may provide complementary information, but its prognostic value for cognitive decline and clinical progression remains unclear.

**Methods:** We studied 1,142 dementia-free participants from the Alzheimer’s Disease Neuroimaging Initiative (ADNI) with available baseline structural MRI, cerebrospinal fluid (CSF) amyloid-β (Aβ42) and phosphorylated tau (p-tau-181), and longitudinal cognitive follow-up. Total hippocampal volume (left + right) and hemispheric asymmetry (absolute left–right volumetric difference) were modeled simultaneously. Linear mixed-effects models examined associations with baseline performance and longitudinal change across memory, language, executive, and visuospatial domains. Cox proportional hazards models assessed risk of clinical progression to clinical dementia over up to 10 years of follow-up. All analyses adjusted for age, sex, education, APOE ε4 status, and CSF biomarkers, with stratification by amyloid status.

**Results:** The study cohort included 546 women (47.8%), with a mean age of 72.54 ± 6.98 years. Larger total hippocampal volume was consistently associated with better baseline performance and slower decline across all four cognitive domains, independent of amyloid and tau biomarkers. In contrast, greater hippocampal asymmetry was selectively associated with worse baseline memory performance and faster memory decline, independent of total hippocampal volume. In amyloid-stratified analyses, total hippocampal volume showed broad associations with cognition across all four domains among amyloid-positive participants and more limited, domain-specific associations among amyloid-negative participants, whereas hippocampal asymmetry was associated with memory only in amyloid-negative individuals. Regarding clinical progression to dementia, smaller total hippocampal volume was associated with higher risk of progression in the overall cohort and within both amyloid groups. In contrast, hippocampal asymmetry was associated with progression risk only among amyloid-negative individuals (hazard ratio per SD increase = 1.31, 95% CI: 1.03–1.65).

**Conclusions:** Hippocampal total volume and asymmetry capture distinct aspects of neurodegeneration, with asymmetry providing additional prognostic information for memory decline and clinical progression in participants without detectable amyloid pathology.

## 1. Introduction

The hippocampus plays a central role in memory and other higher cognitive functions and is among the earliest brain structures affected in Alzheimer’s disease (AD).^1^ Progressive hippocampal atrophy is a well-established neuroimaging biomarker of AD-related neurodegeneration and is strongly associated with cognitive decline and clinical progression to dementia.^2^ However, hippocampal volume loss alone may not fully capture the heterogeneity of cognitive decline observed in older adults, particularly in individuals without overt AD pathology. This has prompted growing interest in additional structural characteristics of the hippocampus that may reflect early or alternative neurodegenerative processes. One such feature is hemispheric asymmetry between the left and right hippocampi, which may indicate imbalanced or focal neurodegeneration not reflected by total volume.^3, 4^ Nevertheless, despite increasing recognition of hippocampal asymmetry in aging and neurodegenerative conditions, it remains unclear whether hippocampal asymmetry provides independent prognostic information for future cognitive decline.

Prior studies have consistently demonstrated associations between hippocampal volume and memory performance across normal aging, mild cognitive impairment (MCI), and AD dementia.^4–7^ However, evidence regarding hemispheric hippocampal asymmetry has been inconsistent, with reported associations varying across populations, study designs, and cognitive outcomes. Some cross-sectional studies have reported greater right-than-left asymmetry is associated with MCI and early AD, while others have found no consistent relationship.^4, 8, 9^ Most existing studies of hippocampal asymmetry have been cross-sectional or restricted to clinically impaired or genetically determined AD populations, limiting inference about its prognostic value. Although asymmetrical hippocampal atrophy has been observed prior to clinical conversion in MCI and autosomal dominant AD,^10, 11^ prognostic value of hippocampal asymmetry in dementia-free older adults remains unclear.

Importantly, hippocampal asymmetry is not specific to AD and has been increasingly linked to other age-related neuropathologic processes that preferentially affect medial temporal structures. Hemispheric differences in hippocampal volume have been reported in association with limbic-predominant age-related TDP-43 encephalopathy (LATE), hippocampal sclerosis of aging, and vascular-related conditions.^12, 13^ LATE, in particular, shows a strong association with hippocampal asymmetry, with higher pathologic stages linked to greater left–right volumetric differences, independent of coexisting AD neuropathology.^14^ These observations suggest that hippocampal asymmetry may reflect focal or asymmetric neurodegeneration arising from mechanisms not primarily driven by amyloid pathology. As a result, asymmetry may capture vulnerability that is not adequately reflected by total hippocampal volume or by biomarkers of core AD pathology. This distinction highlights the need to evaluate hippocampal asymmetry in relation to cognitive decline while explicitly accounting for amyloid and tau burden.

In this study, we examined the independent and complementary contributions of hippocampal volume and hemispheric asymmetry to cognitive decline and clinical progression in dementia-free older adults. Using data from the Alzheimer’s Disease Neuroimaging Initiative (ADNI), we investigated whether baseline hippocampal asymmetry in dementia-free older adults was associated with faster cognitive decline across four cognitive domains: memory, language, executive function, and visuospatial function. By directly modeling hippocampal volume and asymmetry simultaneously and accounting for CSF biomarkers of amyloid and tau, we aimed to distinguish total hippocampal atrophy from hemispheric asymmetry and to evaluate whether these markers differentially reflect neurodegenerative processes related to, or extending beyond, core AD pathology.

## 2. Methods

### 2.1. Participants

#### 2.1.1. ADNI study design

The data used in the preparation of this article were obtained from the ADNI database (adni.loni.usc.edu). The ADNI was launched in 2003 as a public-private partnership, led by Principal Investigator Michael W. Weiner, MD. The primary goal of ADNI has been to test whether serial MRI, PET, other biological markers, and clinical and neuropsychological assessment can be combined to measure the progression of mild cognitive impairment and early dementia. Importantly, ADNI diagnoses are based on standardized clinical criteria and do not require pathological confirmation, allowing for etiologic heterogeneity at the clinical endpoint. ADNI data collection was approved by the institutional review boards of all participating institutions and informed written consent was obtained from all participants. Detailed information on measures and methods of assessment in the ADNI project are available at http://www.adni.loni.usc.edu.

#### 2.1.2. Study participants

We included participants from ADNI (ADNI 1, ADNI GO, ADNI 2, and ADNI 3) who met the following criteria at baseline: (1) availability of high-quality T1-weighted MRI processed through the ADNI FreeSurfer pipeline; (2) availability of baseline CSF concentrations of amyloid-β (Aβ42) and phosphorylated tau at threonine 181 (p-tau-181); and (3) at least two longitudinal cognitive assessments contributing to the harmonized domain composite scores. Participants with a baseline diagnosis of dementia were excluded. After applying these criteria, 1,142 dementia-free individuals were included in the analytic cohort, including 483 cognitively normal (CN) participants and 659 individuals with MCI.

In the ADNI study, diagnostic classification at baseline followed standardized criteria. CN individuals were required to have a Mini-Mental-State Examination (MMSE) scores of 24 or higher; Clinical Dementia Rating (CDR) score of 0, and no evidence of depression. MCI was defined by an MMSE score between 24 and 30 (inclusive); a CDR of 0.5, the presence of memory complaints, and no significant functional impairment. Participants meeting criteria for AD dementia were required to meet the National Institute of Neurological and Communicative Disorders and Stroke–Alzheimer’s Disease and Related Disorders Association (NINCDS-ADRDA) criteria for clinically defined probable AD, an MMSE scores between 20 and 26 (inclusive), and a CDR of 0.5 or 1.^15^ Clinical progression in the present study was defined as meeting ADNI criteria for AD dementia during follow; however, because this outcome reflects a clinical diagnosis rather than biologically confirmed AD, we use the term “dementia” throughout.

### 2.2. Study measures and outcomes

#### 2.2.1. MRI and hippocampal asymmetry

MRI data were automatically processed using the FreeSurfer software package available at http://surfer.nmr.mgh.harvard.edu/) by the Schuff and Tosun laboratory at the University of California-San Francisco as part of the ADNI shared data-set. FreeSurfer methods for identifying and calculation of regional brain volumes have been described in detail previously.^16^

For this study, left (L) and right (R) hippocampal volumes were adjusted for intracranial volume (ICV). Total hippocampal volume was defined as the sum of left and right hippocampal volumes (L+R), and hippocampal asymmetry as the absolute hemispheric difference (|L − R|).

Both total hippocampal volume and hemispheric asymmetry were included simultaneously in all statistical models to distinguish effects attributable to total atrophy from those reflecting asymmetric neurodegeneration. To assess potential collinearity between hippocampal volume and asymmetry, we calculated variance inflation factors (VIFs) and condition indices using a linear regression model including both variables. VIF values were < 5 and condition indices were < 30, indicating no evidence of problematic multicollinearity. Although absolute hemispheric difference may still scale with overall hippocampal volume, the absence of collinearity and the simultaneous modeling of total volume and asymmetry would allow for estimation of independent associations with cognitive and clinical outcomes.

#### 2.2.2. Cognition

Harmonized cognitive domain scores were obtained from the Alzheimer’s Disease Sequencing Project – Phenotype Harmonization Consortium (ADSP-PHC).^17^ The ADSP-PHC performs centralized curation and harmonization of neuropsychological data across multiple aging and dementia cohorts, including ADNI, to improve longitudinal consistency and cross-study comparability. Item-level test data were assigned by expert consensus to four cognitive domains—memory, language, executive function, and visuo-spatial function. These domains were defined a priori based on established neuropsychological frameworks. Cross-cohort “anchor items” (tests administered and scored identically across at least two cohorts) were identified to ensure equivalence of scoring. Confirmatory factor analysis was then used to estimate a harmonized model for each domain, in which anchor item parameters were constrained to be equal across studies while study-specific items were freely estimated. Co-calibrated parameters from these models were subsequently applied to generate composite scores on a common scale (range = −3 to +3) across cohorts and longitudinal time points.^18^

The use of harmonized domain composites, rather than individual neuropsychological tests, was intended to reduce measurement noise, mitigate test-specific practice effects, and enhance sensitivity to longitudinal cognitive change. All cognitive domains were included in analyses a priori, with memory examined as a primary domain of interest based on the known role of the hippocampus in episodic memory.

#### 2.2.3. CSF Biomarkers

CSF samples were batch processed by the ADNI Biomarker Core at the University of Pennsylvania School of Medicine using Roche Elecsys assay.^19^ Baseline CSF p-tau_181_ and Aβ42 levels were included as continuous variables in all models. For descriptive purposes and stratified models, amyloid positivity was defined as CSF Aβ42 < 977 pg/mL, and tau positivity was defined as CSF p-tau181 > 24 pg/mL, consistent with validated ADNI cutpoints.^20^

### 2.3. Statistical analysis

#### Group comparisons

Differences in participant characteristics across sex and age subgroups were tested using independent-sample t tests for continuous variables and Chi Square test for categorical variables.

#### Cognitive trajectories

To evaluate the effect of baseline total hippocampal volume and asymmetry on baseline and longitudinal cognitive performance, we used linear mixed-effects (LME) models. These models accommodate unbalanced follow-up and missing outcome data under a missing-at-random assumption. Time was modeled as a continuous variable representing years since baseline. The base model included fixed effects for time, total hippocampal volume, hippocampal asymmetry, and their interactions with time, as well as covariates (age, sex, years of education, and APOE ε4 status). Random intercepts and slopes for time were included at the participant level to capture individual variability in baseline cognition and rate of cognitive change. Estimates for hippocampal volume and asymmetry reflect their association with baseline cognitive performance (time 0), whereas the corresponding interactions with time represent their association with the rate of cognitive change over time. Separate models were fit for each cognitive domain.

In secondary models, baseline Aβ42 and p-tau were added as covariates in addition to the covariates of the base model. These models were used to evaluate whether associations between hippocampal measures and cognitive outcomes were independent of core AD-related biomarkers. Amyloid-stratified analyses were conducted as secondary, hypothesis-driven analyses to examine potential effect modification by baseline amyloid status. Separate LME models were fit within amyloid-positive and amyloid-negative groups using identical model specifications. Models included fixed effects for time, hippocampal volume, hippocampal asymmetry, and their interactions with time; baseline p-tau as a continuous variable; and covariates of age, sex, education, and APOE ε4 status.

In a separate model, hippocampal asymmetry magnitude was dichotomized into high and low groups based on the 75th percentile of the absolute asymmetry (cutoff value = 0.32 cm^3^). Linear mixed-effects models were used to examine the association of high asymmetry with cognition, adjusting for total hippocampal volume and relevant covariates. The top quartile was selected a priori to identify individuals with pronounced hemispheric differences while maintaining sufficient sample size for stable estimation. Because no validated clinical threshold exists for hippocampal asymmetry, a distribution-based cutoff provides a transparent and reproducible approach for contrasting higher-versus lower-severity asymmetry.

#### Clinical progression (CDR)

Cox proportional hazards models were used to examine whether hippocampal asymmetry predicted risk of clinical progression to dementia. Time to progression was defined as the interval between baseline and the first visit at which a participant met criteria for ADNI dementia diagnosis. Participants who did not progress were censored at their last follow-up visit. Models included age, sex, education, and APOE ε4 carrier status as covariates. Because baseline diagnosis (CN vs. MCI) substantially affects baseline hazard, it was included as a stratification variable rather than a covariate, allowing the baseline hazard to differ between strata while assuming common effects of predictors. Total hippocampal volume and hemispheric asymmetry were entered simultaneously to evaluate independent effects of total volume and asymmetry on progression risk. Kaplan–Meier survival curves stratified by baseline hippocampal asymmetry quartiles were generated for visualization. All statistical analyses were conducted in SPSS version 27 (IBM Corp, Armonk, NY, USA).

## 3. Results

### 3.1. Cohort characteristics

The study cohort consisted of 1,142 dementia-free individuals, of whom 546 (47.8%) were females, and the mean age was 72.54 ± 6.98 years. The average total hippocampal volume in the entire sample was 7.22 ± 1.08 cubic centimeter (cm^3^), and the average hippocampal asymmetry was 0.23 ± 0.17 cm^3^. The right hippocampal was larger than the left hippocampus in the entire cohort (3.67 ± 0.57 vs 3.55 ± 0.54 cm^3^, paired *t*-test, *p* < 0.001). Based on the CSF Aβ42 levels, 577 individuals were classified as amyloid-negative and 565 as amyloid-positive. Mean hippocampal volume was significantly smaller in the amyloid-positive group compared to the amyloid-negative group (6.96 ± 1.07 vs. 7.47 ± 1.03, *p* < 0.001). Hippocampal asymmetry was not different between the amyloid positive and negative groups. Demographic, biomarker, and cognitive domain characteristics of the study cohort are summarized in Table 1.

**Table 1.** Participants’ Characteristics at Baseline.

| Characteristic | All | Aβ42- | Aβ42+ |
| --- | --- | --- | --- |
| N (%) | 1,142 | 577 | 565 |
| Females, n (%) | 546 (47.8) | 296 (51.3) | 250 (44.2) |
| Age, yr | 72.54 (6.98) | 71.73 (7.06) | 73.37 (6.80) |
| Education, yr | 16.26 (2.66) | 16.38 (2.58) | 16.13 (2.73) |
| Aβ42, pg/mL | 1160.67 (634.76) | 1648.35 (534.19) | 662.64 (178.64) |
| P-tau181, pg/mL | 25.02 (12.8) | 20.95 (8.68) | 29.17 (14.84) |
| Hippocampal volume (cm<sup>3</sup>) | 7.22 (1.08) | 7.47 (1.03) | 6.96 (1.07) |
| Hippocampal asymmetry (cm<sup>3</sup>) | 0.23 (0.17) | 0.23 (0.17) | 0.23 (0.17) |
| Memory* | 0.56 (0.64) | 0.78 (0.56) | 0.34 (0.64) |
| Language* | 0.58 (0.53) | 0.71 (0.50) | 0.46 (0.53) |
| Executive function* | 0.53 (0.56) | 0.70 (0.51) | 0.36 (0.56) |
| Visuospatial function* | 0.47 (0.26) | 0.53 (0.49) | 0.41 (0.53) |

Values are presented as mean ± standard deviation (SD) for continuous variables and as number (percentage) for categorical variables. \*Harmonized cognitive domain scores are standardized scores obtained from the Alzheimer's Disease Sequencing Project – Phenotype Harmonization Consortium (ADSP-PHC) and range from -3 to +3. Abbreviations: Aβ42, amyloid-beta 42; p-tau18, phosphorylated tau at threonine 181.

### 3.2. Association of baseline hippocampal asymmetry with cognitive performance

In the base model, larger total hippocampal volume was significantly associated with better baseline cognitive performance across all four cognitive domains (all *p* < 0.001, Table 2). These associations remained significant after adjustment for Aβ, and further for Aβ and tau (all *p* < 0.001). In the base model, greater hippocampal asymmetry was associated with worse performance in memory (β = −0.26 ± 0.10, *p* = 0.009) and language (β = −0.21 ± 0.08, *p* = 0.008); both associations remained significant after adjustment for Aβ and for Aβ plus tau. No significant associations were found between hippocampal asymmetry and executive or visuospatial function in any model.

**Table 2.**
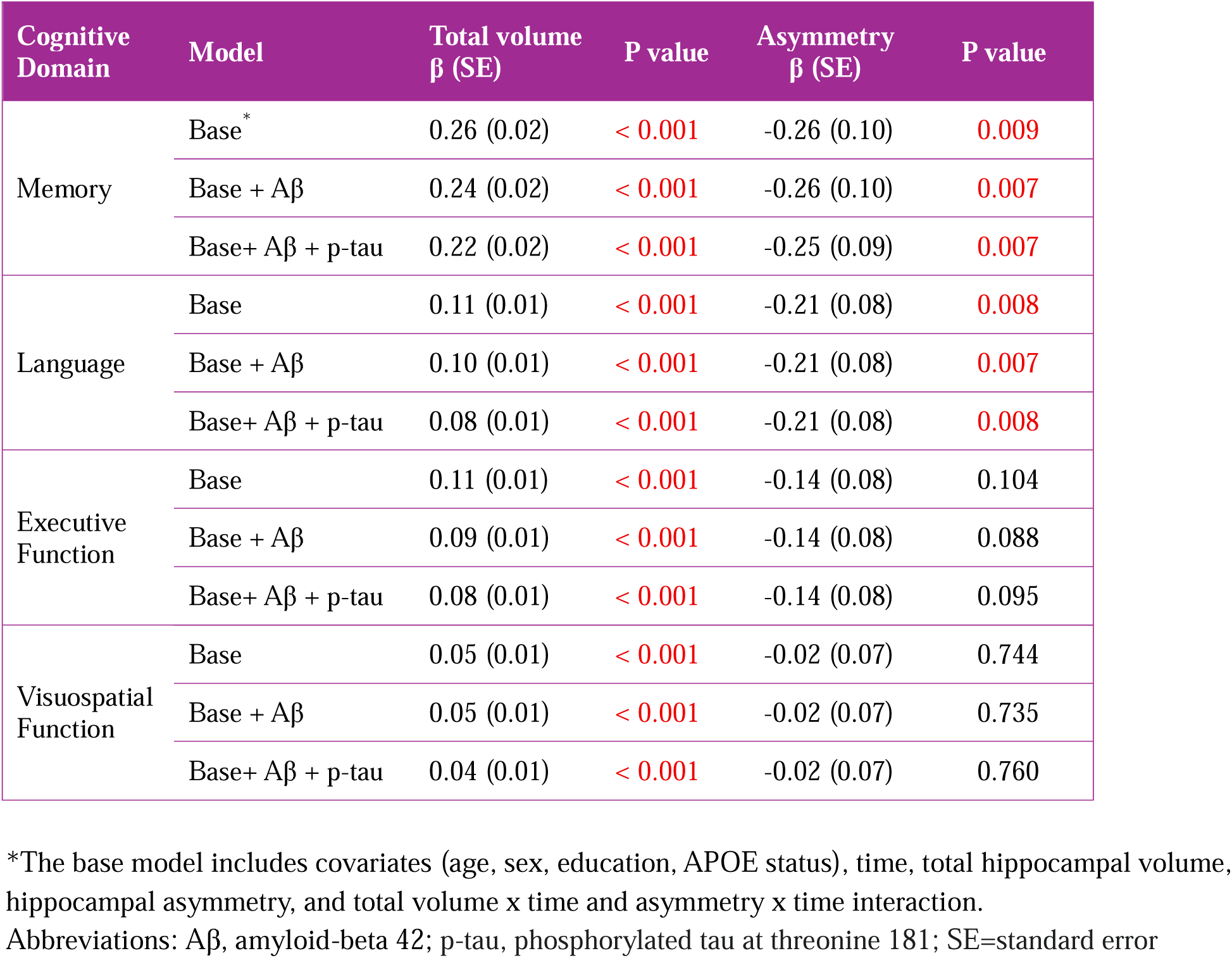
Association of baseline hippocampal total volume and asymmetry with baseline cognition.

In the base model, larger total hippocampal volume was associated with a slower rate of decline across all four cognitive domains (all *p* < 0.001; Table 3). These associations remained significant after adjustment for Aβ, and, subsequently, for Aβ plus tau (both *p* < 0.001). In contrast, greater hippocampal asymmetry predicted a faster rate of decline in memory performance (β = −0.06 ± 0.02, *p* = 0.018). This association remained significant after adjustment for Aβ and for Aβ plus tau (both *p* = 0.018). No significant associations were observed between hippocampal asymmetry and the rate of decline in the other cognitive domains.

**Table 3.** Association of baseline hippocampal total volume and asymmetry with rate of change in cognition.

| Cognitive Domain | Model | Total volume x Time<br>$\beta$ (SE) | P value | Asymmetry x Time<br>$\beta$ (SE) | P value |
| --- | --- | --- | --- | --- | --- |
| Memory | Base* | 0.05 (0.00) | < 0.001 | -0.06 (0.02) | 0.018 |
| | Base + Aβ | 0.05 (0.00) | < 0.001 | -0.06 (0.02) | 0.018 |
| | Base+ Aβ + p-tau | 0.05 (0.00) | < 0.001 | -0.06 (0.02) | 0.018 |
| Language | Base | 0.04 (0.00) | < 0.001 | -0.03 (0.02) | 0.116 |
| | Base + Aβ | 0.04 (0.00) | < 0.001 | -0.03 (0.02) | 0.116 |
| | Base+ Aβ + p-tau | 0.04 (0.00) | < 0.001 | -0.03 (0.02) | 0.116 |
| Executive Function | Base | 0.03 (0.00) | < 0.001 | -0.01 (0.02) | 0.734 |
| | Base + Aβ | 0.03 (0.00) | < 0.001 | -0.01 (0.02) | 0.747 |
| | Base+ Aβ + p-tau | 0.03 (0.00) | < 0.001 | -0.01 (0.02) | 0.756 |
| Visuospatial Function | Base | 0.02 (0.00) | < 0.001 | 0.00 (0.02) | 0.851 |
| | Base + Aβ | 0.02 (0.00) | < 0.001 | 0.00 (0.02) | 0.856 |
| | Base+ Aβ + p-tau | 0.02 (0.00) | < 0.001 | 0.00 (0.02) | 0.857 |

\*The base model includes covariates (age, sex, education, APOE status), time, total hippocampal volume, hippocampal asymmetry, and total volume x time and asymmetry x time interaction.
Abbreviations: Aβ , amyloid-beta 42; p-tau, phosphorylated tau at threonine 181.

To further investigate the effect of amyloid burden, we conducted stratified analyses by dichotomous baseline Aβ status (Table 4). Among Aβ+ individuals, larger total hippocampal volume was associated with higher baseline cognitive scores and a slower rate of decline across all four domains (all *p* < 0.001). In Aβ− participants, larger total hippocampal volume was associated with higher baseline scores and a slower rate of decline in memory and language, as well as higher baseline executive function (Table 4). In contrast, greater hippocampal asymmetry was associated with worse baseline memory performance (β = −0.31 ± 0.12, *p* = 0.013) and faster memory decline (β = −0.07 ± 0.02, *p* < 0.001) only among Aβ− individuals, with no significant associations observed in the Aβ+ group.

**Table 4.**
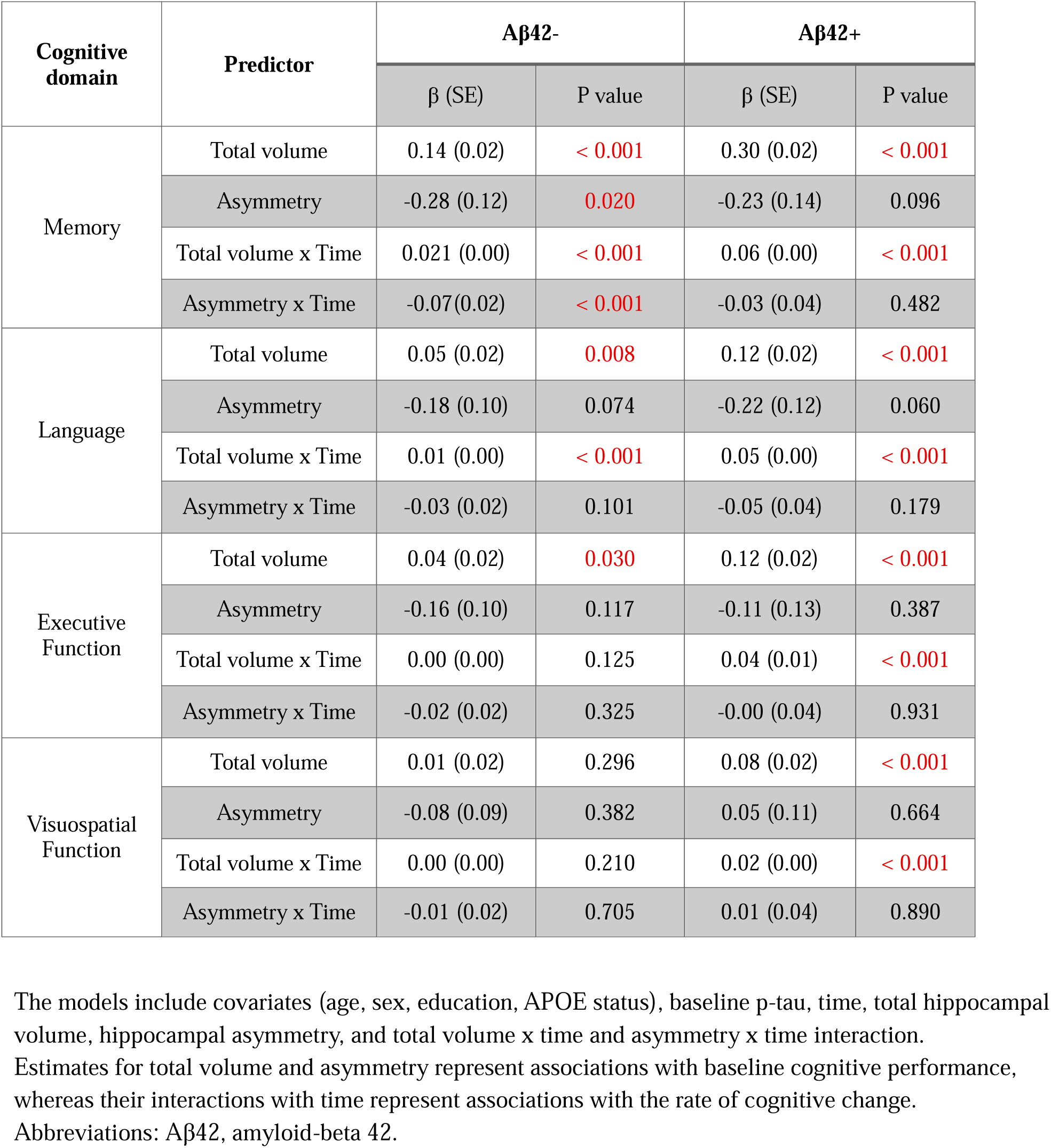
Association of baseline hippocampal total volume and asymmetry with baseline and rate of change in cognition stratified by amyloid status.

| Cognitive domain | Predictor | Aβ42- | | Aβ42+ | |
| --- | --- | --- | --- | --- | --- |
| | | $\beta$ (SE) | P value | $\beta$ (SE) | P value |
| Memory | Total volume | 0.14 (0.02) | < 0.001 | 0.30 (0.02) | < 0.001 |
|  | Asymmetry | -0.28 (0.12) | 0.020 | -0.23 (0.14) | 0.096 |
|  | Total volume x Time | 0.021 (0.00) | < 0.001 | 0.06 (0.00) | < 0.001 |
|  | Asymmetry x Time | -0.07(0.02) | < 0.001 | -0.03 (0.04) | 0.482 |
| Language | Total volume | 0.05 (0.02) | 0.008 | 0.12 (0.02) | < 0.001 |
|  | Asymmetry | -0.18 (0.10) | 0.074 | -0.22 (0.12) | 0.060 |
|  | Total volume x Time | 0.01 (0.00) | < 0.001 | 0.05 (0.00) | < 0.001 |
|  | Asymmetry x Time | -0.03 (0.02) | 0.101 | -0.05 (0.04) | 0.179 |
| Executive Function | Total volume | 0.04 (0.02) | 0.030 | 0.12 (0.02) | < 0.001 |
|  | Asymmetry | -0.16 (0.10) | 0.117 | -0.11 (0.13) | 0.387 |
|  | Total volume x Time | 0.00 (0.00) | 0.125 | 0.04 (0.01) | < 0.001 |
|  | Asymmetry x Time | -0.02 (0.02) | 0.325 | -0.00 (0.04) | 0.931 |
| Visuospatial Function | Total volume | 0.01 (0.02) | 0.296 | 0.08 (0.02) | < 0.001 |
|  | Asymmetry | -0.08 (0.09) | 0.382 | 0.05 (0.11) | 0.664 |
|  | Total volume x Time | 0.00 (0.00) | 0.210 | 0.02 (0.00) | < 0.001 |
|  | Asymmetry x Time | -0.01 (0.02) | 0.705 | 0.01 (0.04) | 0.890 |

The models include covariates (age, sex, education, APOE status), baseline p-tau, time, total hippocampal volume, hippocampal asymmetry, and total volume x time and asymmetry x time interaction.
Estimates for total volume and asymmetry represent associations with baseline cognitive performance, whereas their interactions with time represent associations with the rate of cognitive change.
Abbreviations: Aβ42, amyloid-beta 42.

To determine whether the association between hippocampal asymmetry and memory decline depended on asymmetry direction, we included an interaction term between the asymmetry and laterality (a binary variable defined as L > R vs R > L) in linear mixed-effects models adjusted for the same covariates. The association between hippocampal asymmetry and baseline memory performance did not differ by laterality (|L − R| × laterality, *p* = 0.406), nor did its association with the rate of memory decline (laterality × |L − R| × time, *p* = 0.153), indicating that the effect of asymmetry was independent of hemispheric dominance. The association between hippocampal asymmetry and memory performance remained significant among amyloid negative individuals after accounting for laterality (β = −0.12 ± 0.04, *p* = 0.003).

When hippocampal asymmetry was modeled as a dichotomous variable (high vs. low) rather than as a continuous measure, participants with high asymmetry in the overall cohort did not exhibit significantly worse baseline memory performance (*p* = 0.084) or a faster rate of memory decline compared with those with low asymmetry (*p* = 0.367). However, in amyloid-stratified analyses, high hippocampal asymmetry was significantly associated with faster memory decline among amyloid-negative individuals (β = −0.03 ± 0.00, *p* < 0.001). This association remained significant after accounting for laterality, indicating that the effect of high asymmetry in the amyloid-negative group was independent of hemispheric dominance.

### 3.3. Association of baseline hippocampal asymmetry with risk of clinical progression

Of 1,142 participants included in the study, 259 (22.7%) progressed to a clinical diagnosis of dementia over 10 years. Among amyloid-negative participants, 42 (7.3%) progressed, whereas 217 (38.4%) of amyloid-positive participants progressed.

In the Cox proportional hazards models, smaller baseline hippocampal total volume was associated with a greater risk of progression to dementia (HR = 0.46, 95% CI: 0.40–0.53, *p* = <0.001). In contrast, hippocampal asymmetry was not associated with risk of progression. We then examined the interactive associations between baseline hippocampal asymmetry and amyloid burden on risk of progression to dementia. When baseline Aβ42 and its interaction with hippocampal asymmetry were added to the model, the interaction term was significant (HR = 1.28, 95% CI: 1.00–1.63, *p* = 0.044), indicating that the association between asymmetry and progression risk differs depending on amyloid status. To further investigate the effect of amyloid burden, analyses were stratified by dichotomous Aβ status. Smaller hippocampal volume was associated with increased risk of conversion to dementia in both Aβ positive and negative groups (both *p* <0.001). In contrast, greater hippocampal asymmetry was associated with a higher risk of progression among Aβ− participants (HR = 1.31, 95% CI: 1.03–1.65, *p* = 0.028), but not among Aβ+ participants (HR = 1.04, 95% CI: 0.91–1.20, *p* = 0.529; Fig 1). This hazard ratio indicates a 31% higher risk of progression per one standard deviation increase in hippocampal asymmetry in Aβ− individuals.

**Figure 1.**
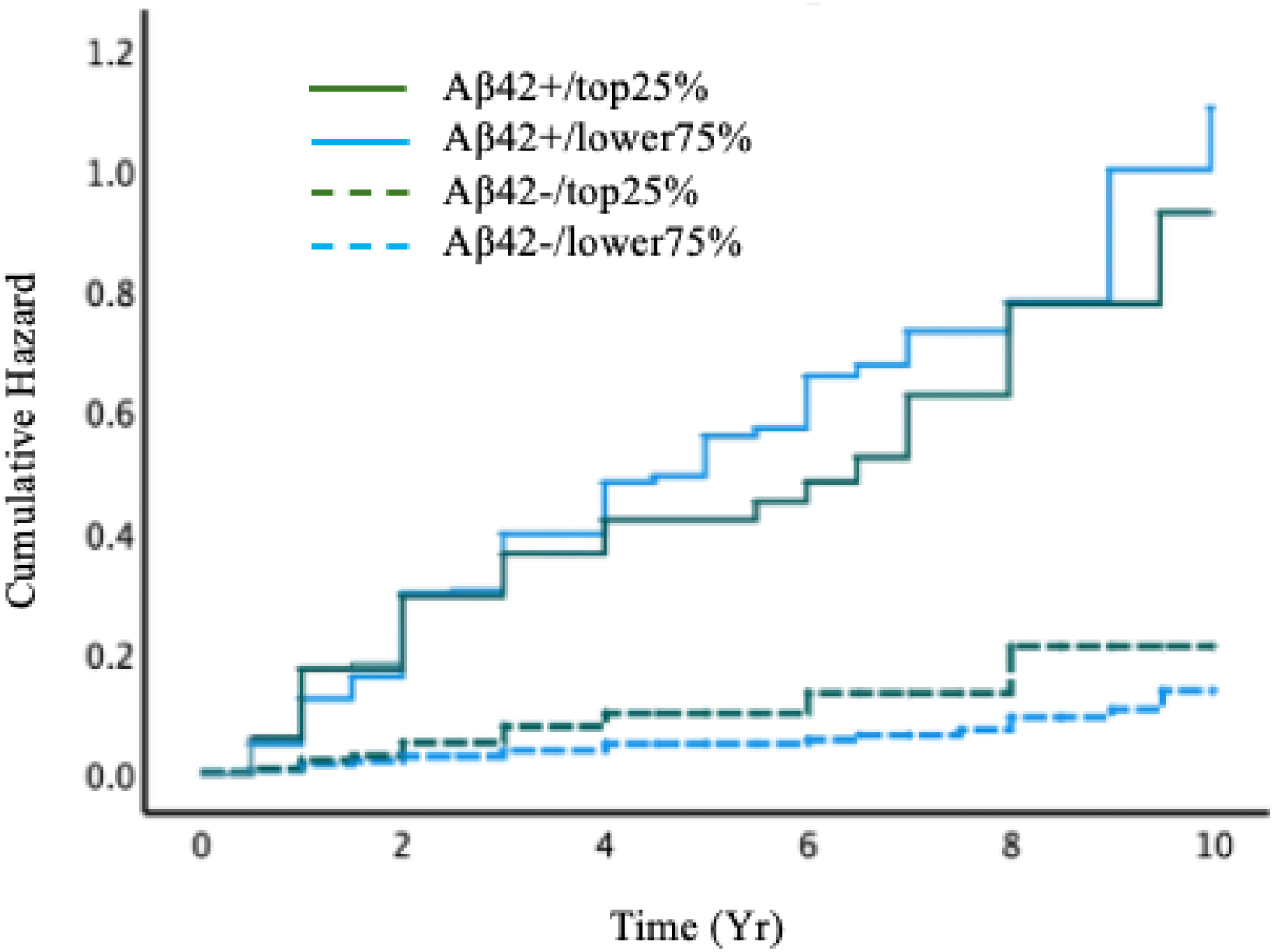
Cumulative hazard of progression to dementia by baseline hippocampal asymmetry (top quartile vs. others) stratified by baseline amyloid status

## 4. Discussion

Our findings highlight complementary and independent roles of hippocampal total volume and asymmetry in capturing cognitive decline. While hippocampal total volume was consistently associated with baseline performance and longitudinal change across all four cognitive domains, hemispheric asymmetry showed a more selective association with memory, independent of total hippocampal volume and after adjustment for Aβ and tau. When comparing Aβ− and Aβ+ individuals, hippocampal total volume was significantly smaller in the amyloid-positive group; however, hippocampal asymmetry did not differ significantly between the two groups. Amyloid-stratified analyses further demonstrated that total hippocampal volume was consistently associated with cognitive performance across all four domains among Aβ+ individuals and with fewer domains in Aβ− individuals, whereas hippocampal asymmetry was specifically associated with memory performance only in Aβ− participants. Similarly, smaller total hippocampal volume was associated with a higher risk of clinical progression to dementia in the overall cohort and within both amyloid groups; in contrast, hippocampal asymmetry was associated with clinical progression only among Aβ− individuals.

Together, these findings suggest that while total hippocampal degeneration is aligned with broader cognitive vulnerability, hemispheric asymmetry may capture focal or lateralized degeneration that is most apparent in the memory domain. Furthermore, the finding that asymmetry effects were primarily observed in Aβ− participants suggests the possibility that asymmetry reflects etiologic processes distinct from core amyloid-related pathways.

Previous studies have reported mixed findings regarding hippocampal asymmetry in aging and dementia, with some showing greater right- or left-lateralized atrophy in AD and others finding no consistent pattern.^4^ A key challenge in interpreting this literature is that asymmetry has been operationalized using heterogeneous metrics, and many studies have not modeled asymmetry jointly with total hippocampal volume. The present study extends prior work by modeling hippocampal total volume and asymmetry as separate, simultaneous predictors, allowing a clearer distinction between global hippocampal atrophy and left-right volumetric imbalance.

Our findings on the association between total hippocampal volume and cogitation align well with the existing literature. Cross-sectional studies have shown that larger hippocampal volumes are associated with better episodic and working memory in cognitively normal older adults.^21^ In individuals with MCI and AD, smaller hippocampal volumes have been strongly linked to lower cognitive performance.^22^ Longitudinal analyses further support that smaller baseline hippocampus and greater atrophy in specific subfields predict subsequent cognitive decline.^23–25^ In contrast, the relationship between hippocampal asymmetry and cognition remains less clear. While some studies report that greater asymmetry, especially in subfields such as CA1 or the dentate gyrus, is associated with poorer memory and learning in MCI and AD, findings in healthy older adults have generally shown little or no association between asymmetry and cognitive performance.^7, 21, 26, 27^ Consistent with this pattern of heterogeneity, our results indicate that hippocampal total volume shows broad associations with cognition, whereas asymmetry shows more selective associations, primarily in memory, after accounting for total volume.

Notably, the observed association between hippocampal asymmetry and memory performance was independent of the core AD pathologies. This suggests that the left-right volumetric imbalance may reflect neurodegenerative processes not fully captured by amyloid and tau biomarkers, potentially including LATE, hippocampal sclerosis or vascular pathologies.^28–30^ Consistent with this interpretation, amyloid-stratified analyses indicated that asymmetry was associated with memory performance only among Aβ− individuals, whereas hippocampal volume remained more broadly associated with cognition among Aβ+ participants. Together, these findings support a model in which total hippocampal atrophy is more tightly coupled to AD-spectrum neurodegeneration, while hemispheric asymmetry may serve as a complementary marker of focal degeneration that is most evident when amyloid burden is low.

Similarly, our finding that smaller hippocampal total volume was associated with increased risk of progression to dementia in the entire cohort is strongly supported by prior research. Several longitudinal and meta-analytic studies have demonstrated that hippocampal atrophy is a robust predictor of conversion to AD in both CN and MCI individuals.^31–34^ Studies comparing MCI converters with stable MCI participants have shown that medial temporal neurodegeneration is the most reliable marker of progression, with reduced hippocampal volume emerging as the most robust predictor.^10^ In our combined cohort of CN and MCI individuals, hippocampal asymmetry was not associated with risk of conversion in the overall sample. However, in amyloid-stratified analyses, asymmetry predicted progression to dementia only among Aβ− participants. This pattern further supports the notion that hippocampal asymmetry may be particularly informative for identifying both memory decline and clinical progression among individuals without evidence of amyloid pathology.

A major strength of the present study lies in our analytic approach, which simultaneously modeled hippocampal total volume and hemispheric asymmetry. Prior studies examining hippocampal asymmetry have often relied on the asymmetry index, defined as (L−R)/(L+R). However, this metric can introduce scaling artifacts when total hippocampal volume differs across study groups. For example, in our sample, total hippocampal volume (L+R) was significantly larger in CN compared to MCI, and the absolute hemispheric difference (|L−R|) did not differ between groups. Nonetheless, the asymmetry index appeared significantly larger in MCI than CN solely because the denominator (L+R) was smaller in MCI, not because of greater true left–right differences. Thus, the elevated asymmetry index in MCI reflects a mathematical artifact rather than a biological phenomenon. To avoid this issue, we modeled total hippocampal volume and hemispheric difference as separate variables, allowing a clearer distinction between effects attributable to overall atrophy versus true hemispheric asymmetry and enabling a more reliable interpretation of their associations with cognitive and clinical outcomes. Our study also has limitations. We analyzed a single cohort and did not replicate the findings in an independent dataset. The relatively high educational attainment and predominantly White composition of the ADNI sample may limit the generalizability of these findings to more diverse populations. In addition, hippocampal volume and asymmetry were assessed only at baseline; longitudinal assessment of structural changes may provide greater sensitivity for detecting early neurodegenerative processes.

In conclusion, our findings suggest that, beyond overall hippocampal volume, greater left–right volumetric differences may serve as an early imaging marker of memory decline and clinical progression, particularly among individuals without detectable amyloid pathology. Validation of these results in independent cohorts is needed to establish the clinical utility of hippocampal asymmetry as a prognostic imaging biomarker.

## Data Availability

All data produced in the present study are available upon reasonable request to the authors

## Acknowledgment

Data collection and sharing for this project was funded by the Alzheimer’s Disease Neuroimaging Initiative (ADNI) (National Institutes of Health Grant U01 AG024904) and DOD ADNI (Department of Defense award number W81XWH-12-2-0012). ADNI is funded by the National Institute on Aging, the National Institute of Biomedical Imaging and Bioengineering, and through generous contributions from the following: AbbVie, Alzheimer’s Association; Alzheimer’s Drug Discovery Foundation; Araclon Biotech; BioClinica, Inc.; Biogen; Bristol-Myers Squibb Company; CereSpir, Inc.; Cogstate; Eisai Inc.; Elan Pharmaceuticals, Inc.; Eli Lilly and Company; EuroImmun; F. Hoffmann-La Roche Ltd and its affiliated company Genentech, Inc.; Fujirebio; GE Healthcare; IXICO Ltd.; Janssen Alzheimer Immunotherapy Research & Development, LLC.; Johnson & Johnson Pharmaceutical Research & Development LLC.; Lumosity; Lundbeck; Merck & Co., Inc.; Meso Scale Diagnostics, LLC.; NeuroRx Research; Neurotrack Technologies; Novartis Pharmaceuticals Corporation; Pfizer Inc.; Piramal Imaging; Servier; Takeda Pharmaceutical Company; and Transition Therapeutics. The Canadian Institutes of Health Research is providing funds to support ADNI clinical sites in Canada. Private sector contributions are facilitated by the Foundation for the National Institutes of Health (www.fnih.org). The grantee organization is the Northern California Institute for Research and Education, and the study is coordinated by the Alzheimer’s Therapeutic Research Institute at the University of Southern California. ADNI data are disseminated by the Laboratory for Neuro Imaging at the University of Southern California.

The ADSP Phenotype Harmonization Consortium (ADSP-PHC) is funded by NIA (U24 AG074855, U01 AG068057 and R01 AG059716). The harmonized cohorts within the ADSP-PHC include:C:the Anti-Amyloid Treatment in Asymptomatic Alzheimer’s study (A4 Study), a secondary prevention trial in preclinical Alzheimer’s disease, aiming to slow cognitive decline associated with brain amyloid accumulation in clinically normal older individuals. The A4 Study is funded by a public-private-philanthropic partnership, including funding from the National Institutes of Health-National Institute on Aging, Eli Lilly and Company, Alzheimer’s Association, Accelerating Medicines Partnership, GHR Foundation, an anonymous foundation and additional private donors, with in-kind support from Avid and Cogstate. The companion observational Longitudinal Evaluation of Amyloid Risk and Neurodegeneration (LEARN) Study is funded by the Alzheimer’s Association and GHR Foundation. The A4 and LEARN Studies are led by Dr. Reisa Sperling at Brigham and Women’s Hospital, Harvard Medical School and Dr. Paul Aisen at the Alzheimer’s Therapeutic Research Institute (ATRI), University of Southern California. The A4 and LEARN Studies are coordinated by ATRI at the University of Southern California, and the data are made available through the Laboratory for Neuro Imaging at the University of Southern California. The participants screening for the A4 Study provided permission to share their de-identified data to advance the quest to find a successful treatment for Alzheimer’s disease. We would like to acknowledge the dedication of all the participants, the site personnel, and all of the partnership team members who continue to make the A4 and LEARN Studies possible. The complete A4 Study Team list is available on: a4study.org<u>/a4-study-team.</u>; the Adult Changes in Thought study (ACT), *U01 AG006781, U19 AG066567;* Alzheimer’s Disease Neuroimaging Initiative (ADNI): *Data collection and sharing for this project was funded by the Alzheimer’s Disease Neuroimaging Initiative (ADNI) (National Institutes of Health Grant U01 AG024904) and DOD ADNI (Department of Defense award number W81XWH-12-2-0012). ADNI is funded by the National Institute on Aging, the National Institute of Biomedical Imaging and Bioengineering, and through generous contributions from the following: AbbVie, Alzheimer’s Association; Alzheimer’s Drug Discovery Foundation; Araclon Biotech; BioClinica, Inc.; Biogen; Bristol-Myers Squibb Company; CereSpir, Inc.; Cogstate; Eisai Inc.; Elan Pharmaceuticals, Inc.; Eli Lilly and Company; EuroImmun; F. Hoffmann-La Roche Ltd and its affiliated company Genentech, Inc.; Fujirebio; GE Healthcare; IXICO Ltd.;Janssen Alzheimer Immunotherapy Research & Development, LLC.; Johnson & Johnson Pharmaceutical Research & Development LLC.; Lumosity; Lundbeck; Merck & Co., Inc.;Meso Scale Diagnostics, LLC.; NeuroRx Research; Neurotrack Technologies; Novartis Pharmaceuticals Corporation; Pfizer Inc.; Piramal Imaging; Servier; Takeda Pharmaceutical Company; and Transition Therapeutics. The Canadian Institutes of Health Research is providing funds to support ADNI clinical sites in Canada. Private sector contributions are facilitated by the Foundation for the National Institutes of Health (*www.fnih.org*). The grantee organization is the Northern California Institute for Research and Education, and the study is coordinated by the Alzheimer’s Therapeutic Research Institute at the University of Southern California. ADNI data are disseminated by the Laboratory for Neuro Imaging at the University of Southern California;* Estudio Familiar de Influencia Genetica en Alzheimer (EFIGA): *5R37AG015473, RF1AG015473, R56AG051876;* the Health & Aging Brain Study – Health Disparities (HABS-HD), supported by the National Institute on Aging of the National Institutes of Health under Award Numbers R01AG054073, R01AG058533, R01AG070862, P41EB015922, and U19AG078109; the Korean Brain Aging Study for the Early Diagnosis and Prediction of Alzheimer’s disease (KBASE), which was supported by a grant from Ministry of Science, ICT and Future Planning (Grant No: NRF-2014M3C7A1046042); Memory & Aging Project at Knight Alzheimer’s Disease Research Center (MAP at Knight ADRC): The Memory and Aging Project at the Knight-ADRC (Knight-ADRC). This work was supported by the National Institutes of Health (NIH) grants R01AG064614, R01AG044546, RF1AG053303, RF1AG058501, U01AG058922 and R01AG064877 to Carlos Cruchaga. The recruitment and clinical characterization of research participants at Washington University was supported by NIH grants P30AG066444, P01AG03991, and P01AG026276. Data collection and sharing for this project was supported by NIH grants RF1AG054080, P30AG066462, R01AG064614 and U01AG052410. We thank the contributors who collected samples used in this study, as well as patients and their families, whose help and participation made this work possible. This work was supported by access to equipment made possible by the Hope Center for Neurological Disorders, the Neurogenomics and Informatics Center (NGI: https://neurogenomics.wustl.edu/) and the Departments of Neurology and Psychiatry at Washington University School of Medicine; National Alzheimer’s Coordinating Center (NACC): *The NACC database is funded by NIA/NIH Grant U24 AG072122.* SCAN is a multi-institutional project that was funded as a U24 grant (AG067418) by the National Institute on Aging in May 2020. Data collected by SCAN and shared by NACC are contributed by the NIA-funded ADRCs as follows*: P30 AG062429 (PI James Brewer, MD, PhD), P30 AG066468 (PI Oscar Lopez, MD), P30 AG062421 (PI Bradley Hyman, MD, PhD), P30 AG066509 (PI Thomas Grabowski, MD), P30 AG066514 (PI Mary Sano, PhD), P30 AG066530 (PI Helena Chui, MD), P30 AG066507 (PI Marilyn Albert, PhD), P30 AG066444 (PI John Morris, MD), P30 AG066518 (PI Jeffrey Kaye, MD), P30 AG066512 (PI Thomas Wisniewski, MD), P30 AG066462 (PI Scott Small, MD), P30 AG072979 (PI David Wolk, MD), P30 AG072972 (PI Charles DeCarli, MD), P30 AG072976 (PI Andrew Saykin, PsyD), P30 AG072975 (PI David Bennett, MD), P30 AG072978 (PI Neil Kowall, MD), P30 AG072977 (PI Robert Vassar, PhD), P30 AG066519 (PI Frank LaFerla, PhD), P30 AG062677 (PI Ronald Petersen, MD, PhD), P30 AG079280 (PI Eric Reiman, MD), P30 AG062422 (PI Gil Rabinovici, MD), P30 AG066511 (PI Allan Levey, MD, PhD), P30 AG072946 (PI Linda Van Eldik, PhD), P30 AG062715 (PI Sanjay Asthana, MD, FRCP), P30 AG072973 (PI Russell Swerdlow, MD), P30 AG066506 (PI Todd Golde, MD, PhD), P30 AG066508 (PI Stephen Strittmatter, MD, PhD), P30 AG066515 (PI Victor Henderson, MD, MS), P30 AG072947 (PI Suzanne Craft, PhD), P30 AG072931 (PI Henry Paulson, MD, PhD), P30 AG066546 (PI Sudha Seshadri, MD), P20 AG068024 (PI Erik Roberson, MD, PhD), P20 AG068053 (PI Justin Miller, PhD), P20 AG068077 (PI Gary Rosenberg, MD), P20 AG068082 (PI Angela Jefferson, PhD), P30 AG072958 (PI Heather Whitson, MD), P30 AG072959 (PI James Leverenz, MD);* National Institute on Aging Alzheimer’s Disease Family Based Study (NIA-AD FBS): U24 AG056270; Religious Orders Study (ROS): P30AG10161,R01AG15819, R01AG42210; Memory and Aging Project (MAP - Rush): R01AG017917, R01AG42210; Minority Aging Research Study (MARS): R01AG22018, R01AG42210; the Texas Alzheimer’s Research and Care Consortium (TARCC), funded by the Darrell K Royal Texas Alzheimer’s Initiative, directed by the Texas Council on Alzheimer’s Disease and Related Disorders; Washington Heights/Inwood Columbia Aging Project (WHICAP): *RF1 AG054023;*and Wisconsin Registry for Alzheimer’s Prevention (WRAP): *R01AG027161 and R01AG054047.* Additional acknowledgments include the National Institute on Aging Genetics of Alzheimer’s Disease Data Storage Site (NIAGADS, U24AG041689) at the University of Pennsylvania, funded by NIA.

